# Using geographic information systems to link population estimates to wastewater surveillance data in New York State, USA

**DOI:** 10.1101/2022.08.23.22279124

**Authors:** Dustin T. Hill, David A. Larsen

**Author notes:** Correspondence to:* Dustin T. Hill.

## Abstract

Sewer systems provide many services to communities that have access to them beyond removal of waste and wastewater. Understanding of these systems’ geographic coverage is essential for wastewater-based epidemiology (WBE), which requires accurate estimates for the population contributing wastewater. Reliable estimates for the boundaries of a sewer service area or sewershed can be used to link upstream populations to wastewater samples taken at treatment plants or other locations within a sewer system. These geographic data are usually managed by public utilities, municipal offices, and some government agencies, however, there are no centralized databases for geographic information on sewer systems. We created a database for all municipal sewersheds in New York State for the purpose of supporting statewide wastewater surveillance efforts to support public health. We used a combination of public tax records with sewer access information, physical maps, and municipal records to organize and draw digital boundaries compatible with geographic information systems. The methods we employed to create these data will be useful to inform similar efforts in other jurisdictions and the data have many public health applications as well as being informative for water/environmental research and infrastructure projects.

## 1 Introduction

Wastewater from a community provides a wealth of information about that population including types of infectious diseases circulating, types of drugs consumed, and types of food eaten (1,2). Wastewater-based epidemiology (WBE) is the use of wastewater to understand public health issues through testing wastewater samples and then estimating community-level trends (1,2). WBE became exponentially important during the emergence of COVID-19 with governments using wastewater surveillance to track the SARS-CoV-2 virus to inform disease spread, intensity, and genetic variation (3). Beyond COVID-19, WBE has a strong record of informing our understanding of antimicrobial resistance (4), expanding the study of endemic diseases, such as influenza and HIV, and guiding public health research and policy into the future (5). In addition to providing information on infectious disease threats to public health (6), testing wastewater can inform understanding of the burden of opioid and other drug use in a community (7). Linking the population to the wastewater sample is a key step in WBE (8,9), as accurate population estimates are needed to understand what is found in wastewater in terms of population-level burden and create statistical models that use these data (10). Linking the sample to the population is also necessary for precise public health intervention if an outbreak is detected (11).

The area from which a treatment plant receives wastewater is known as the sewershed, defined as an area of land where all the sewers flow into one point (12). Sewersheds are usually polygon shapes inclusive of the property parcels that have connections to the sewer system but can also include larger portions of land area that may contribute runoff rainwater to the system (13). Sewersheds follow natural hydrology (like rivers and streams), flowing downhill congruent with topography in most cases. There are, however, sewer systems that “jump” rivers with pipes crossing the banks aboveground or going underneath the waterway. Systems can also include what are known as force mains, which are sewer lines that force water uphill to the final collection system (14). This variation complicates the estimation of boundaries for sewer systems when survey maps do not exist, but there are ways to approximate boundaries such as using municipal borders and tax registries that collect information on whether a parcel of land is connected to public sewer or not (15). The sewershed boundary can also be linked with other geographically-referenced data, such as population census data, to estimate the population within a sewershed (16), and linkages can be made with environmental data, such as precipitation and temperature, to better understand wastewater test results. Linking wastewater samples to geographically relevant data can enhance research on infectious disease transmission and improve the utility of data related to the social determinants of health.

Geographic data on sewer systems, including manhole locations, sewer lines, and related infrastructure, are usually maintained by the municipality that owns the system or the engineers that designed it originally (DEC, personal communication). In other words, centralized databases for sewers across different jurisdictions are uncommon and likely unavailable (NY GIS, personal communication). As New York State’s wastewater surveillance network increased in scale and scope, a centralized database for all of New York State’s municipal sewer systems was needed to link testing of municipal wastewater to upstream populations for public health surveillance of SARS-CoV-2, other pathogens, and substances of public health interest. Since no centralized database existed beyond a list of the point locations for each treatment plant per discharge permitting records (17), creation of a database of sewersheds was necessary geographic data for the state’s wastewater surveillance network. The final product is a spatial database that has every municipal sewer system in New York State mapped and linked to the treatment plant and population served by that sewer system. In what follows, we detail the methods for gathering and drawing the final sewershed polygons, and steps for data validation and quality assurance. These methods can be applied in other jurisdictions with similar needs and we explore some of the uses for these date beyond public health.

## 2 Data and Methods

### 2.1 Data sources

#### 2.1.1 Treatment plant operators

The NY Department of Environmental Conservation (DEC) provided a database listing 638 municipal wastewater treatment plants (17). This list included the name of the facility, its location via street address and geocoordinates, as well as other metadata such as permitted discharge capacity. The list of these plants were the targets for this project with the goal being to find and draw sewershed boundaries for each plant. Contact information for 385 of the plants on the list was provided by the DEC, and a survey was distributed to these plants via email and phone in 2021 (see Hill et al., 2022 for the full question list).

The purpose of the survey was to determine what data the treatment plant operators had regarding the service area of the facility and what they knew about WBE methods for tracking substances in wastewater for public health. Results on operator knowledge regarding WBE has been published previously (18). If no data existed, the operator provided a description of the service area to facilitate drawing of the boundary using other data sources. The respondents were followed up with email and phone calls to request they send the data for inclusion in the database. Data were received in different formats including digital shapefiles of the boundary, sewer manholes, and/or sewer mains; physical maps of the boundary, sewer manholes, and/or sewer mains; and address lists of the properties connected to the sewer system.

#### 2.1.2 New York State Parcel database

NY maintains a geodatabase of property tax data for all properties in the state. While data for several counties is publicly available, most data are not publicly accessible. We received permission from the state Geographic Information Systems office for use of the data to aid in construction of the sewershed boundaries. The tax parcel data consisted of spatial polygons and included a field indicating if the parcel was on public sewer or a private system. This allowed classification of parcels based on sewer type. In addition, special district tables were provided that included information on water and sewer districts that could be joined to the tax parcels by parcel ID number. The sewer district data was not complete for all counties, but, for counties where it was available, it was used to classify separate sewer systems that were adjacent. The version of the data used was from 2020 (19).

#### 2.1.3 Municipal websites and other data sources

In addition to calling municipalities, we researched public sewer systems online through town, village, and city websites searching for any published records, reports, or information on the location of sewer service areas. From public websites, we were able to obtain copies of physical maps and descriptions of some sewer systems. A list of websites for source material is provided in the supplementary material.

For the three counties in northern New York, additional data were provided by the Development Authority of the North Country (DANC), which is a public benefit corporation set up to manage the infrastructure needs for those counties (20).

DANC provided estimates for 55 sewershed boundaries based on work they completed using village boundaries, sewer billing data, and sewer district data. Further description is available in the supplemental material.

### 2.2 Combination of data

Data were collected and compiled for each county in NY using eight methods listed in Table 1. Wastewater treatment plants (WWTPs) were mapped to the county to provide the initial locations for communities that would have access to public sewer. The first sewersheds that were added were those that were provided by the treatment plant or municipal/county officials and these were not modified. Following these, if we had an address list for properties that were billed for public sewer by the municipality, we matched parcels from the NY tax parcel data for the county. The boundaries for the parcels were then dissolved to form the final boundary.

**Table 1:**
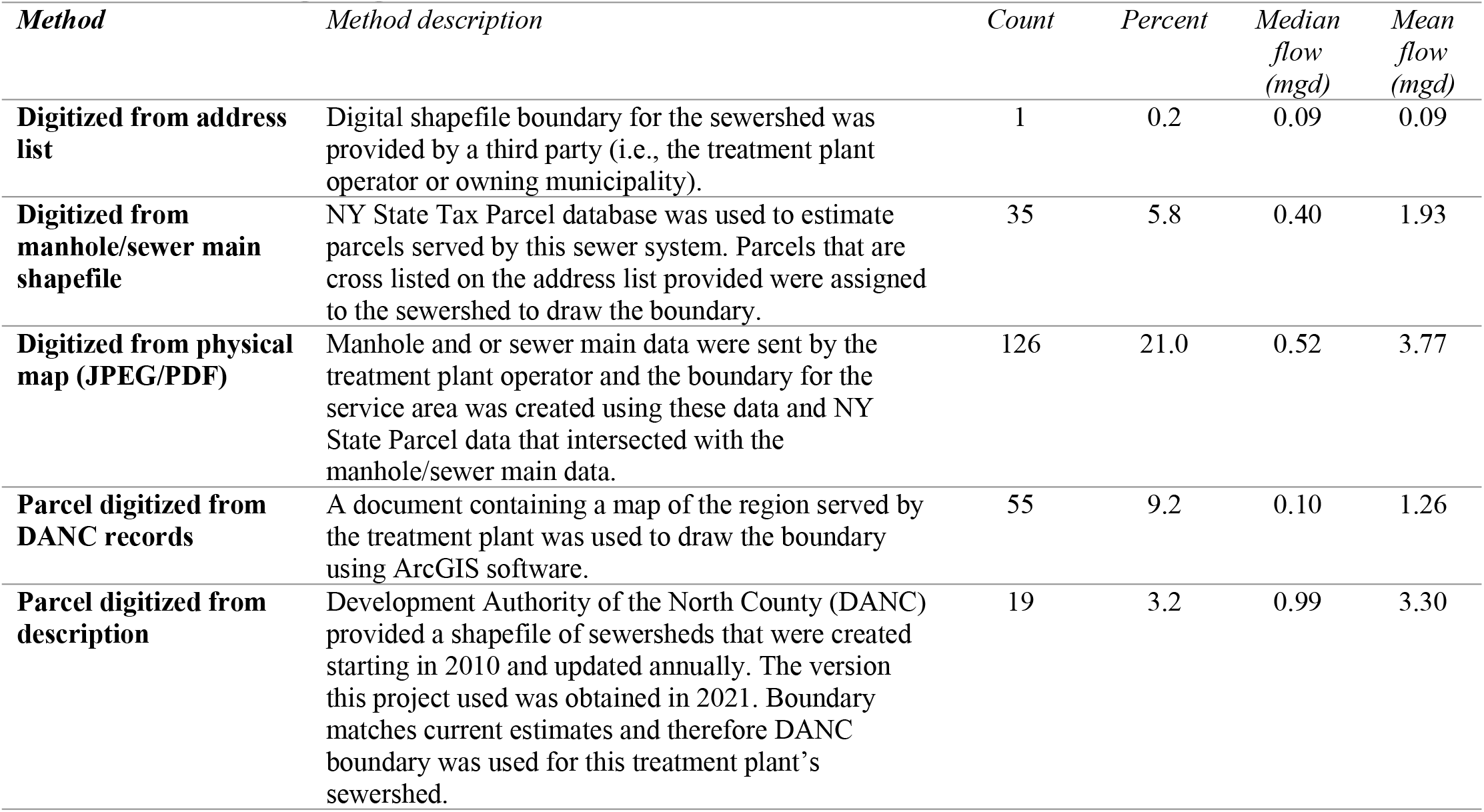
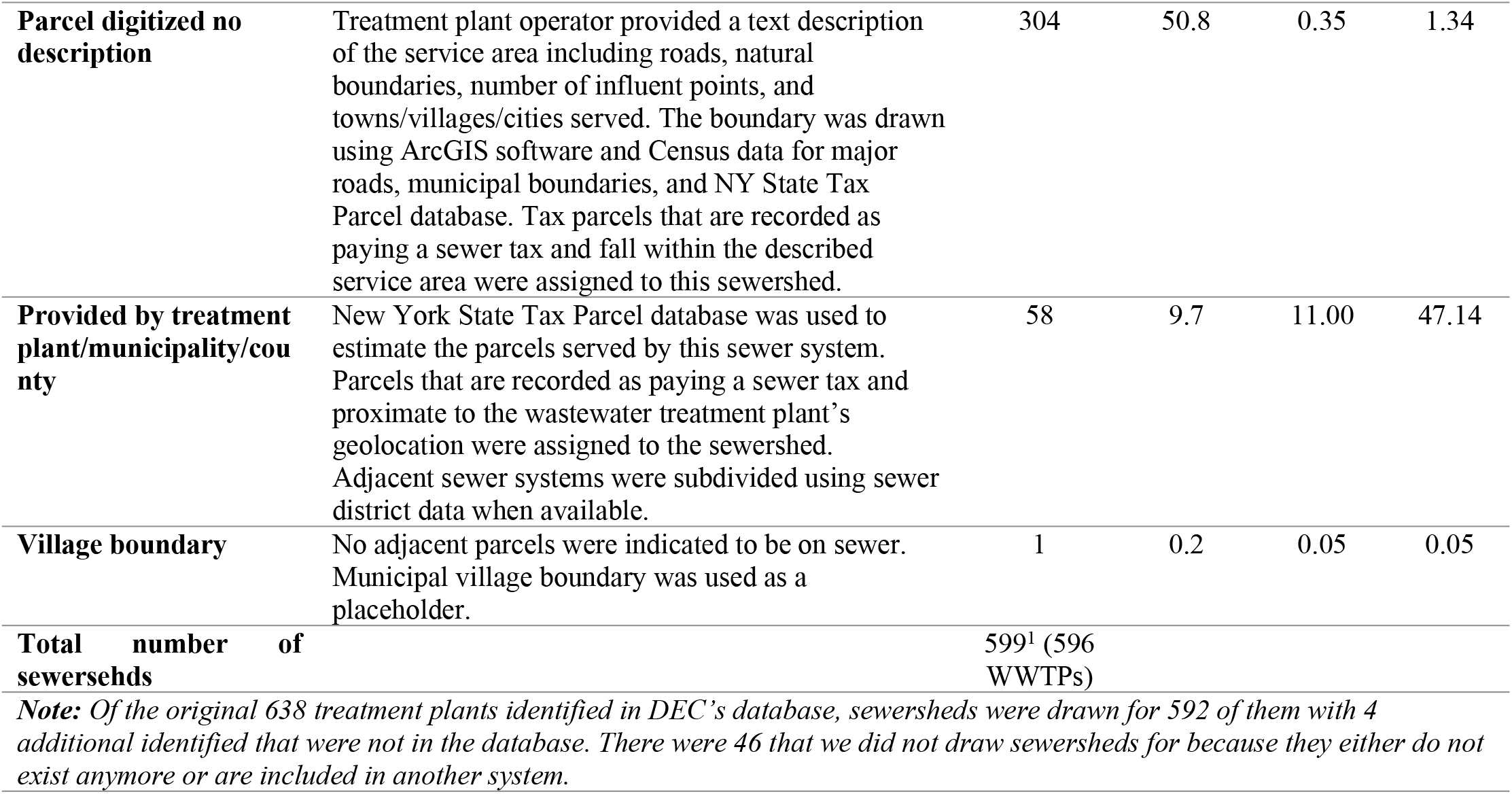
Methods for digitizing sewersheds.

For municipalities that we received digital manhole or sewer main data, we mapped those against the tax parcel data for the county and selected the parcels that were adjacent to sewer mains and proximate to manhole locations (Fig. 1a and Fig. 1b). These parcels were then dissolved to form the sewershed boundary and linked to the municipality’s treatment plant. The other type of data provided were physical maps showing the boundaries of the service area. For these maps, we aligned the borders of the map with the tax parcel data for that county and municipality and then selected the parcels within the boundary (Fig. 1c). Listed roads and other landmarks that formed the boundary were used to ensure the correct parcels were selected. The parcel boundaries were then dissolved to form the final sewershed boundary.

**Figure 1:**
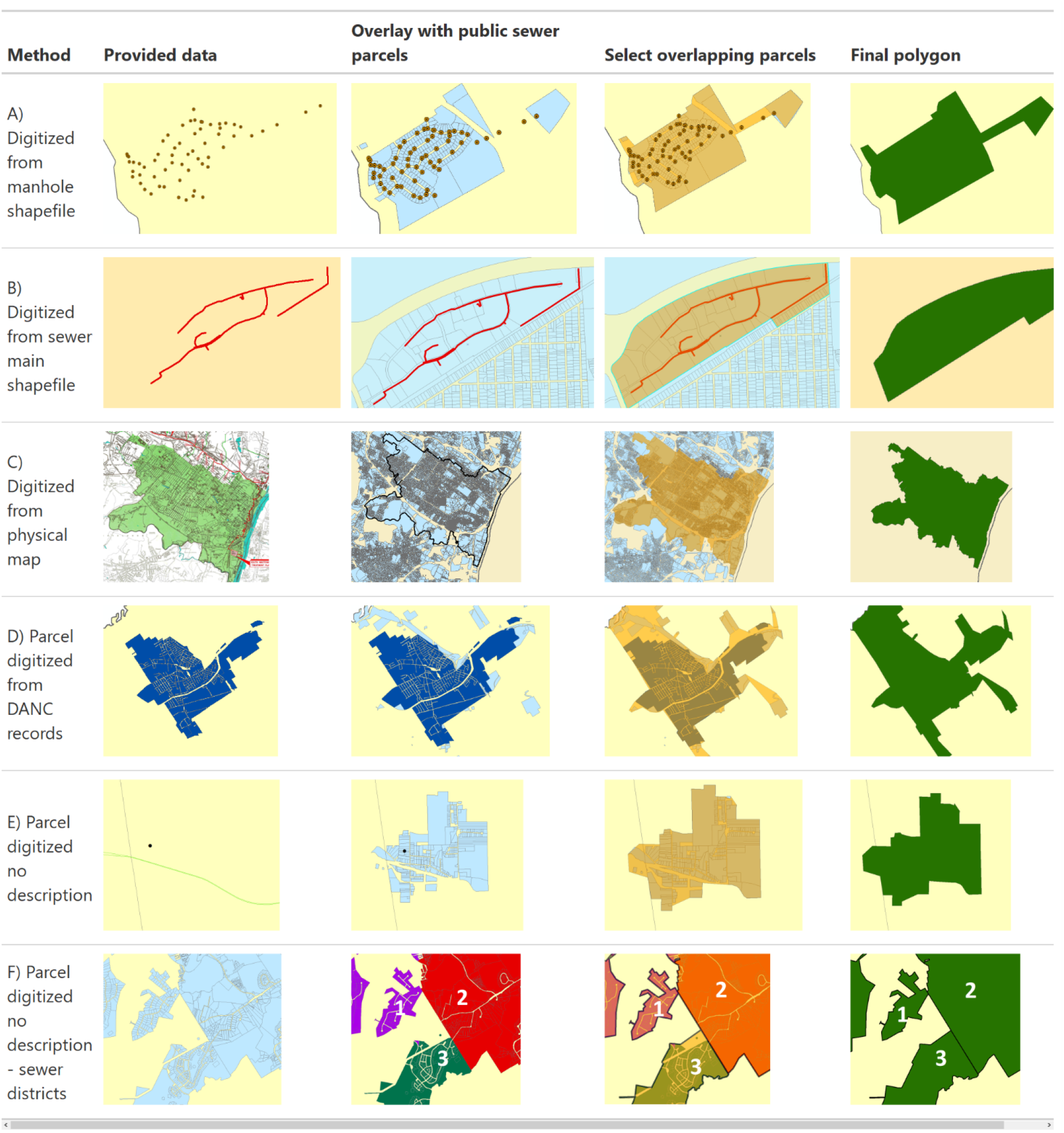
Methods used to create sewersheds.

Boundaries provided by DANC were compared to the reported parcels on public sewer. For many municipalities, the boundaries corresponded well and the DANC boundary was used, however, in some cases adjacent parcels were reported to be on public sewer but not in the DANC sewershed. Therefore, we added these additional parcels to form the final sewershed combining the DANC sewershed with adjacent parcels on public sewer for the same municipality (Fig. 1d).

For the remaining municipalities that we did not have data provided or could not find public records for sewer boundaries or districts, we drew the boundaries based on the parcels on public sewer near the geolocation of the municipal WWTP (Fig. 1e). For many locations, the clusters of sewered parcels corresponded well to the locations of treatment plants and there were clear boundaries between sewer systems. For some counties with several WWTPs serving adjacent communities, it was harder to distinguish between the sewer parcels and the correct WWTP to link it to. For those locations, we examined the sewer district data from the tax parcel special district tables. Where available, sewer districts were identified and grouped together to form distinct sewer systems that were then linked to the correct WWTP (Fig. 1f). Some counties did not have good sewer district data and for those the treatment plant or municipal official was contacted to request information on the boundaries for adjacent sewer systems.

### 2.3 Adding population data

Estimates for 2020 population by sewershed were calculated using R statistical software version 4.1.1 (21). We first estimated the 2010 population for each sewershed using an overlay of 2010 U.S. Census blocks on top of the sewershed boundaries. Census blocks were selected because they represent the smallest geometry for public census data and increasing the accuracy for the intersection with sewershed boundaries. We calculated the proportion of the area for partial block overlap and then assigned the proportional 2010 decennial population of the block to the sewershed assuming equal distribution of the population in the blocks. The apportioned values were then aggregated to obtain a total population estimate for the sewershed.

Then, we repeated this procedure using 2010 decennial population data for the block group and 2018 American Community Survey (ACS) data for the block group to get 2010 and 2018 population by sewershed based on block groups. We used these values to estimate the annual rate of population change per sewershed using the following formula:

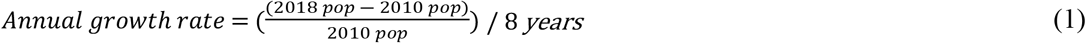

We then applied this average annual change to the sewershed population based on the block data from 2010 and estimated the population after ten years of growth using the following formula to calculate 2020 population estimates:

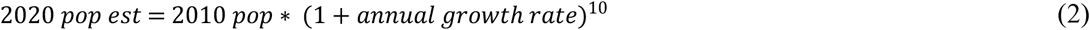

We obtained all population estimates using the “tidycensus” R package (22) and we use the “tigris” package (23) for U.S. Census geometry data. R code for US census intersections is publicly available at https://github.com/dthill196/NY-Sewershed-Populations.

### 2.4 Validation and quality assurance

We validated sewershed boundaries by cross referencing data types when multiple forms were available such as having physical maps and shapefiles. For sewersheds where the boundaries were in question or more difficult to estimate, direct contact and consultation with treatment plant operators was used to check boundaries against the knowledge of the treatment plant staff. Short of formal land surveys of the municipality, the data we created are not easily validated. To check accuracy for all sewersheds, we ran correlations of the population estimates, population density, and surface area of the final polygons with the permitted discharge capacity. Discharge capacity increases with population served and was one way to check that methods for creating sewersheds were producing good estimates for areas served, and the method has been used by other researchers working on population and treatment plant linkages (24).

## 3 Results

We received 116 completed surveys with 19 responses that had a description of the service area. These were then combined with other data sources, such as the NY Tax Parcel database, to draw the sewershed boundary. The remaining 97 responses indicated that the plant had data on the service area of the plant. The survey respondent was then contacted requesting they share the data with our research team. We received digital data for the boundaries of 58 treatment plants and digital data for manholes and sewer mains for 35 treatment plants, and an address list for one treatment plant. Data were retrieved in a combination of survey call-backs and contact with treatment plants after the survey ended (Table 1). In total, 294 sewersheds were digitized using data provided or descriptions of the system representing 49.1 percent of the final database for NY sewersheds and these facilities were among the largest in the state with a median flow rate of 11 millions of gallons per day (mgd) and mean of 47 mgd (Table 1).

The remaining sewersheds (304) were digitized from NY tax parcel data and sewer district data with one sewershed digitized using the village municipal boundary (Table 1). Of the original 638 municipal treatment plants identified using the DEC database, we were able to create boundaries for 592 of them. There were 46 WWTPs in the DEC list that we did not create sewershed boundaries for because they either did not exist anymore or were additional permits for coincident treatment plants representing combined sewer overflow facilities or large pump stations (see supplemental material for a list of permits and reasons they were excluded). In addition, we created sewershed boundaries for 4 additional WWTPs not in the DEC database. Two of these facilities are owned by Native American communities and are on land owned by those communities with their permits supervised by Federal agencies. We included them because their boundaries were evident from tax parcel data for public sewer that were not linked to any treatment plant in the DEC list. Two additional facilities were reported by county governments with their permit information and physical maps provided showing their locations. The final count for our database is 596 municipal WWTP sewersheds. Three WWTPs have two sewersheds each representing separate influent streams from different parts of their service area bringing the total number of sewersheds to 599 (Table 1).

The treatment plants are distributed around the state with some regions and counties having many separate sewer systems with others having fewer (Fig. 2). The total population in the state estimated to be on public sewer is over 16 million, which is approximately 82.6 percent of the total state population (using 19.5 million as the estimated NY population in 2020). Population coverage per sewershed ranged between less than 100 people to as high as 1.2 million. Population density ranged between 1.6 people per km^2^ and 28,000 people per km^2^. Surface area ranged between 58,000 m^2^ and 990 km^2^. There was high correlation between the permitted discharge capacity of each facility and estimated population served (Pearson correlation = 0.897, Fig. 3), moderately high correlation between discharge capacity and estimated population density (Pearson correlation = 0.569, Fig. 3), and high correlation between discharge capacity and surface area (Pearson correlation =0.852, Fig. 3). When examining correlations across the different methods for creating sewersheds (Table 2), we find high all Pearson correlation values higher than 0.5 and statistically significant (p-value < 0.05) except for one method. The use of manhole and sewer main data was correlated significantly with population and surface area, but for population density, we found lower correlation with a Pearson estimate of 0.195 and p-value = 0.27 (Table 2).

**Table 2:** Correlation results for each method.

| **Method** | *Discharge capacity and*<br>*estimated population* |  | *Discharge capacity and*<br>*population density* |  | *Discharge capacity and*<br>*surface area* |  |
| --- | --- | --- | --- | --- | --- | --- |
|  | *Pearson* | *p-* | *Pearson* | *p-value* | *Pearson* | *p-value* |
|  | *correlation* | *value* | *correlation* |  | *correlation* |  |
| **Provided by treatment**<br>**plant/municipality/county** | 0.966 | <0.01 | 0.675 | <0.01 | 0.673 | <0.01 |
| **Parcel digitized no description** | 0.83 | <0.01 | 0.439 | <0.01 | 0.826 | <0.01 |
| **Parcel digitized from**<br>**description** | 0.974 | <0.01 | 0.615 | <0.01 | 0.853 | <0.01 |
| **Digitized from physical map**<br>**(JPEG/PDF)** | 0.876 | <0.01 | 0.515 | <0.01 | 0.854 | <0.01 |
| **Digitized from manhole/sewer**<br>**main shapefile** | 0.707 | <0.01 | 0.195 | 0.27 | 0.668 | <0.01 |
| **Parcel digitized from DANC**<br>**records** | 0.907 | <0.01 | 0.444 | 0.001 | 0.842 | <0.01 |

**Figure 2:**
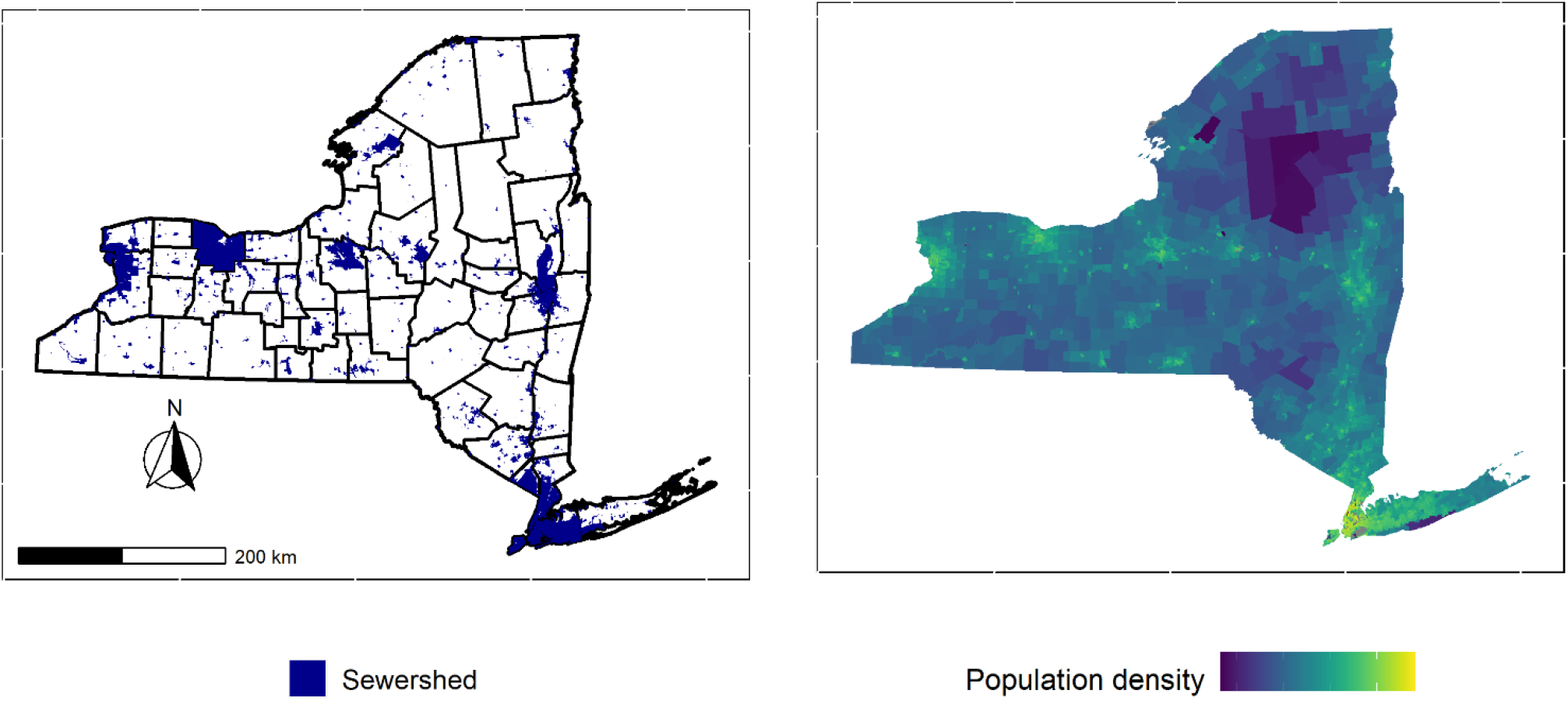
Spatial distribution of sewersheds in New York. Highly sewered counties and regions correspond with population centers across the state.

**Figure 3:**
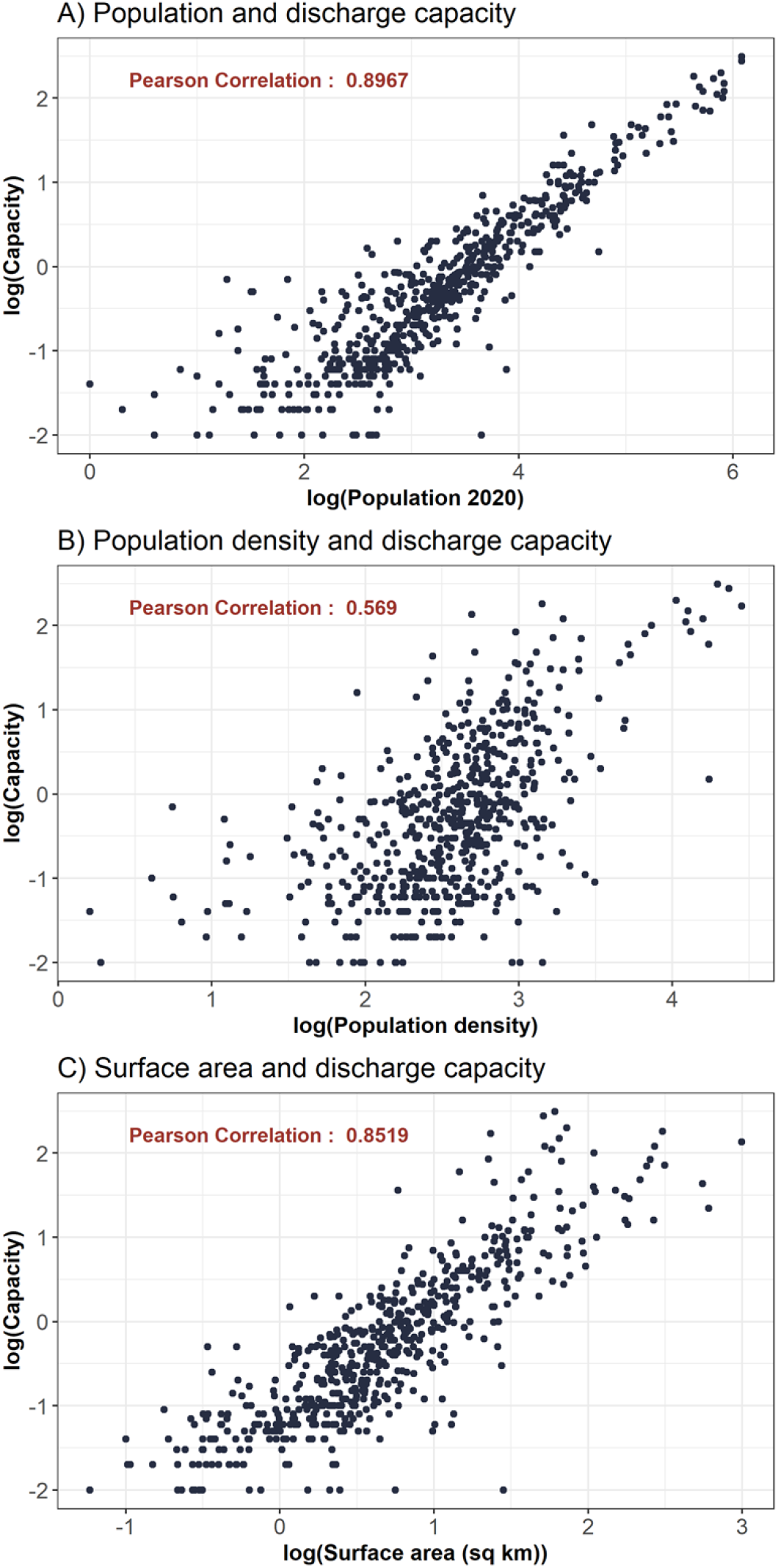
Correlations between permitted discharge capacity of WWTPs and A) estimated population, B) population density, and C) surface area of the sewershed.

## 4 Data Availability

Data described in this manuscript is available upon request to the author. Links to publicly available databases used in this project are provided in the reference section and supplemental documents. R code for creating US census intersections is publicly accessible at https://github.com/dthill196/NY-Sewershed-Populations.

## 5 Discussion and Conclusions

### 5.1 Evaluation of sewershed boundaries

Correlation results between the final sewershed polygons, the discharge capacity of the WWTPs, and the three metrics of population, population density, and surface area were high for all methods used (Figure 3) suggesting that each method contributed acceptable results. One exception to this were sewersheds drawn using manhole or sewer main shapefile data (Table 2). This method resulted in the lowest correlation between discharge capacity and population density, however, there was high correlation for estimated population and surface area for this method. Therefore, despite the one low value for this method, we feel that the resulting sewershed boundaries are acceptable estimates for the service areas of the treatment plants. There were 35 sewersheds drawn using this method with the majority (n = 21) from one county. Removing the county in question from the correlation test yielded a new correlation coefficient of 0.671 (p-value = 0.012) suggesting there might be greater uncertainty with the sewershed boundaries from that county.

The county in question is unique in being highly populated but most of the county population not living on public sewer and, instead of the more typical private septic system for residents, most of the public wastewater goes into what are known as cesspools (25). The county in question has a population over 1 million people but our estimates report only 397,000 of the residents are on public sewer meaning it has high population density but low public sewer population, which might lead to the lower correlation between population density and discharge capacity of the sewer systems. In addition, the county has 30 WWTPs with 22 plants with discharge capacities below 1 million gallons per day. Therefore, the county might be an anomaly among counties in NY with regards to public sewer systems. Despite the low correlation for population density, the county has high correlation for population estimates and surface area with discharge capacity suggesting the resulting boundaries are comparable in their estimates to the rest of the state.

The resulting database of sewersheds includes 4 treatment plants not in the DEC database. These additional plants were drawn based on information from the municipalities and because there were public sewer parcels that matched the description by the municipal officials. DEC’s public database could be behind in its updates between discharge permit documents and adding locations with the most recent evaluation of sewer infrastructure from 2008 (26). We also did not create boundaries for 46 treatment plants listed in DEC’s database for several reasons including that they do not exist anymore or they are included in the sewershed of another facility because they are a smaller pump station or overflow facility. A complete list of the facilities not included and explanations why are provided in the supplemental material.

### 5.2 Applications for public health, research, and policy

The NY Sewershed database was created to support public health efforts related to WBE including the important role of providing population estimates for wastewater test results (10,16). Linking wastewater results to the upstream population is essential for interpretation of findings (6) particularly for case and incidence data for health outcomes of concern such as COVID-19 (27), Polio (28) and other enteroviruses, and opioid use (29). Community-based research that uses wastewater data needs accurate estimates for population served by the facility and large-scale investigations that need to scale up quickly may not have the time to create boundaries or provide estimates in a timely manner particularly if the data are to be used for public health policy. Linking wastewater surveillance data to communities is necessary for policy applications of WBE with increasing application in the US since the start of the COVID-19 pandemic (3,30). Beyond WBE, this database could be used to support water quality research, hydrology research, and policy related to infrastructure improvements.

Water quality researchers might find utility in these data examining freshwater systems and discharge locations. With the sewershed boundaries, researchers now have estimates for where sewer systems border or even cross waterways enabling inclusion of these data in research plans as has been done before (9). In addition, hydrology research can use these data to understand the flow of wastewater to treatment plants from communities and potential relationships with topography and hydrography (31). Lastly, the infrastructure needed to build and maintain sewer systems is aging in NY according the DEC (26) with 36.2 billion dollars estimated to repair, replace, or update sewer systems. Comprehensive mapping of the served communities through the NY sewershed database might be helpful in state-wide and county-wide planning for consolidating sewer systems. While not an official survey of sewered communities or systems, the database we created provides an important step in identifying current sewer systems in NY.

### 5.3 Limitations and future directions

The sewersheds that we created come with some limitations. For all data created, there is uncertainty around the estimated boundaries because they were not produced from formal surveys, but instead methods that estimate the boundary based on secondary data sources. Physical maps that were used might be the most accurate followed by digital data provided by treatment plants, however, the date of some of the maps’ original drawing go back several years, sometimes to the original building of the treatment plant. In addition, the tax parcel data are up-to-date but come with limitations of their own including disclaimers regarding their accuracy. Therefore, these data should not be used in place of official surveys or assessments of sewer systems for the purpose of projects related to infrastructure updates. Instead, these data can support where surveys could be done and help locate sewer systems if previously unmapped.

Further, US Census data estimates represent the population at one time point and data may not be the best match for some locations that are tourist destinations or have frequently population changes due to seasonal residents. Locations of this kind should use the provided population estimates adjusted for their research endeavor and where necessary seek better estimates from other sources. In addition, low population estimates were obtained for many locations (56 sewersheds had population estimates below 100). These low population levels are due to the limits built into the US Census data where population numbers may be suppressed due to low number of residents in rural communities. Similar to sewersheds with population fluctuation, alternate sources may be needed to improve estimation for small sewersheds such as seeking information directly from municipalities on number of sewer hookups.

The future of these data will require updates as time progresses with the potential for sewer systems to be consolidated and new systems to be built making current estimates less reliable. Updates can be done using revised tax parcel data showing additional parcels on sewer as well as surveys of treatment plant operators and municipalities to identify any changes. Methods described in this paper can be applied in other jurisdictions including surveying treatment plants for data availability and working with public and private repositories of land use and property data to estimate Sewershed boundaries. In addition, these methods can be used to aid in formulation of upstream and community-level sampling approaches for public health where sewersheds are sub-divided into smaller units to better identify outbreaks. The potential for changes in sewer infrastructure is a potential challenge, but the current dataset and methods are valuable in their comprehensiveness, novelty, and utility to support public health, water research, and policy.

## Data Availability

Data from public sources is provided in the supplemental material. Data created as part of this project is available upon request to the authors. Data have not been published to a repository because they are owned by several different jurisdictions with some having restricted data access.

https://github.com/dthill196/NY-Sewershed-Populations

## Acknowledgements

This project was made possible by the CDC’s Environmental Public Health and Emergency Response Program, NYS Unique Federal Award Number NUE1EH001341 (NYS Environmental Public Health Tracking Network Maintenance and Enhancement to Accommodate Sub-County Indicators). We would also like to thank Ed Hampston, AJ Smith, and Eric Weigert for their contributions regarding WWTP locations and contact information. Also, a thank-you to Hannah Cousins, Bryan Dandaraw, Catherine Faruolo, Alex Godinez, Sythong Run, Simon Smith, Megan Willkens, and Shruti Zirath for their help calling treatment plants. Additional thanks to Kate Kiyanitsu from the NY GIS office for providing access and support for NY tax parcel data. Thank-you to Tabassum Insaf, Abigail Stamm, and Jeff Bryant from NY DOH for helping refine some Sewershed boundaries. Finally, thank-you to Star Carter at the Development Authority of the North Country for providing estimates for municipal sewer districts for several treatment plants.

## References

1. Lorenzo M, Picó Y. Wastewater-based epidemiology: current status and future prospects. Curr Opin Environ Sci Health. 2019 Jun 1;9:77–84.

2. Sims N, Kasprzyk-Hordern B. Future perspectives of wastewater-based epidemiology: Monitoring infectious disease spread and resistance to the community level. Environ Int. 2020 Jun 1;139:105689.

3. Kirby AE, Walters MS, Jennings WC, Fugitt R, LaCross N, Mattioli M, et al. Using Wastewater Surveillance Data to Support the COVID-19 Response — United States, 2020–2021. Morb Mortal Wkly Rep. 2021 Sep 10;70(36):1242–4.

4. Chau K, Barker L, Budgell E, Vihta K doris, Sims N, Kasprzyk-Hordern B, et al. Systematic Review of Wastewater Surveillance of Antimicrobial Resistance in Human Populations. 2021 Jun 21 [cited 2021 Jul 26]; Available from: https://www.preprints.org/manuscript/202010.0267/v2

5. Kilaru P, Hill D, Anderson K, Collins MB, Green H, Kmush BL, et al. Wastewater surveillance for infectious disease: a systematic review. Am J Epidemiol. In press; 2021.07.26.21261155.

6. Melvin RG, Hendrickson EN, Chaudhry N, Georgewill O, Freese R, Schacker TW, et al. A novel wastewater-based epidemiology indexing method predicts SARS-CoV-2 disease prevalence across treatment facilities in metropolitan and regional populations. Sci Rep. 2021 Nov 1;11(1):21368.

7. Boogaerts T, Ahmed F, Choi PhilM, Tscharke B, O’Brien J, De Loof H, et al. Current and future perspectives for wastewater-based epidemiology as a monitoring tool for pharmaceutical use. Sci Total Environ. 2021 Oct 1;789:148047.

8. Li Z, Li X, Porter D, Zhang J, Jiang Y, Olatosi B, et al. Monitoring the Spatial Spread of COVID-19 and Effectiveness of Control Measures Through Human Movement Data: Proposal for a Predictive Model Using Big Data Analytics. JMIR Res Protoc. 2020 Dec 18;9(12):e24432.

9. Nelson JR, Lu A, Maestre JP, Palmer EJ, Jarma D, Kinney KA, et al. Space-time analysis of COVID-19 cases and SARS-CoV-2 wastewater loading: A geodemographic perspective. Spat Spatio-Temporal Epidemiol. 2022 Aug 1;42:100521.

10. Hoar C, Li Y, Silverman AI. Assessment of Commonly Measured Wastewater Parameters to Estimate Sewershed Populations for Use in Wastewater-Based Epidemiology: Insights into Population Dynamics in New York City during the COVID-19 Pandemic. ACS EST Water [Internet]. 2022 Apr 20 [cited 2022 Jun 28]; Available from: https://doi.org/10.1021/acsestwater.2c00052

11. Xagoraraki I, O’Brien E. Wastewater-Based Epidemiology for Early Detection of Viral Outbreaks. In: O’Bannon DJ, editor. Women in Water Quality: Investigations by Prominent Female Engineers [Internet]. Cham: Springer International Publishing; 2020 [cited 2022 Jun 28]. p. 75–97. (Women in Engineering and Science). Available from: https://doi.org/10.1007/978-3-030-17819-2_5

12. Basaldua J. Watersheds and Sewersheds [Internet]. 2020 [cited 2022 Jun 28]. Available from: https://urbanecologycenter.org/item/1386-watersheds-and-sewersheds.html

13. Sun N, Hong B, Hall M. Assessment of the SWMM model uncertainties within the generalized likelihood uncertainty estimation (GLUE) framework for a high-resolution urban sewershed. Hydrol Process. 2014;28(6):3018–34.

14. Selvakumar A, Tafuri AN, Morrison R, Sterling R. State of technology for renewal of sewer force mains. Urban Water J. 2011 Sep 1;8(5):279–92.

15. Manson SM, Sander HA, Ghosh D, Oakes JM, Orfield MW Jr, Craig WJ, et al. Parcel Data for Research and Policy. Geogr Compass. 2009;3(2):698–726.

16. Tscharke BJ, O’Brien JW, Ort C, Grant S, Gerber C, Bade R, et al. Harnessing the Power of the Census: Characterizing Wastewater Treatment Plant Catchment Populations for Wastewater-Based Epidemiology. Environ Sci Technol. 2019 Sep 3;53(17):10303–11.

17. DEC. Wastewater Treatment Plants | State of New York [Internet]. 2022 [cited 2022 Jun 28]. Available from: https://data.ny.gov/Energy-Environment/Wastewater-Treatment-Plants/2v6p-juki

18. Hill DT, Cousins H, Dandaraw B, Faruolo C, Godinez A, Run S, et al. Wastewater treatment plant operators report high capacity to support wastewater surveillance for COVID-19 across New York State, USA. Sci Total Environ. 2022 Sep 1;837:155664.

19. NY GIS. NYS GIS - Parcels [Internet]. 2022 [cited 2022 Jun 28]. Available from: http://gis.ny.gov/parcels/

20. DANC. Danc - Development Authority of the North Country - About [Internet]. 2022 [cited 2022 Jun 28]. Available from: https://www.danc.org/about

21. R Core Team. R: A language and environment for statistical computing. [Internet]. Vienna, Austria: R Foundation for Statistical Computing; 2021. Available from: https://www.R-project.org/

22. Kyle Walker, Herman M. tidycensus: Load US Census Boundary and Attribute Data as “tidyverse” and ‘sf’-Ready Data Frames [Internet]. 2022. Available from: https://CRAN.R-project.org/package=tidycensus

23. Walker K. tigris: Load Census TIGER/Line Shapefiles [Internet]. 2022. Available from: https://CRAN.R-project.org/package=tigris

24. Ehalt Macedo H, Lehner B, Nicell J, Grill G, Li J, Limtong A, et al. Distribution and characteristics of wastewater treatment plants within the global river network. Earth Syst Sci Data. 2022 Feb 9;14(2):559–77.

25. DEC. Coastal Resiliency and Water Quality in Nassau and Suffolk Counties. 2021;24.

26. DEC. Wastewater Infrastructure Needs of New York State Report - NYS Dept. of Environmental Conservation [Internet]. 2008 [cited 2022 Jun 28]. Available from: https://www.dec.ny.gov/chemical/42383.html

27. Nagarkar M, Keely SP, Jahne M, Wheaton E, Hart C, Smith B, et al. SARS-CoV-2 monitoring at three sewersheds of different scales and complexity demonstrates distinctive relationships between wastewater measurements and COVID-19 case data. Sci Total Environ. 2021 Nov 13;151534.

28. Link-Gelles R. Public Health Response to a Case of Paralytic Poliomyelitis in an Unvaccinated Person and Detection of Poliovirus in Wastewater — New York, June–August 2022. MMWR Morb Mortal Wkly Rep [Internet]. 2022 [cited 2022 Aug 19];71. Available from: https://www.cdc.gov/mmwr/volumes/71/wr/mm7133e2.htm

29. Wang S, C. Green H, L. Wilder m, Du Q, L. Kmush B B. Collins M, et al. High-throughput wastewater analysis for substance use assessment in central New York during the COVID-19 pandemic. Environ Sci Process Impacts. 2020;22(11):2147–61.

30. Reeves K, Liebig J, Feula A, Saldi T, Lasda E, Johnson W, et al. High-resolution within-sewer SARS-CoV-2 surveillance facilitates informed intervention. Water Res. 2021 Oct 1;204:117613.

31. Shuster WD, Bonta J, Thurston H, Warnemuende E, Smith DR. Impacts of impervious surface on watershed hydrology: A review. Urban Water J. 2005 Dec 1;2(4):263–75.

